# Impact of transitioning to newer utility on sex-specific quality-adjusted life expectancy in Norway

**DOI:** 10.1101/2025.09.13.25335693

**Authors:** Mohammad Sayeef Alam

**Affiliations:** Department of Public Health and Nursing, Faculty of Medicine and Health Sciences, Norwegian University of Science and Technology, Trondheim, Norway

**Author notes:** Corresponding author: Mohammad Sayeef Alam.

**Keywords:** EQ-5D-5L, EQ-5D-3L, QALY, prioritization, Norway, UK Crosswalk

## Abstract

Norway has mandated the adoption of the new Norwegian EQ-5D-5L value set from September 1, 2025. This transition change was long overdue, as the former value set was based on decades old assessment of health-related quality of life.

Current study disseminates the updated quality-adjusted life expectancies (QALE) for the Norwegian population based on age and sex, and quantify how the new norm alters the absolute and proportional shortfall in QALY.

The population norm derived from the general population survey by Garratt et al., 2021 has been linked to the most recent Norwegian Lifetable (2023-2024) to estimate the QALE using the Sullivan method. It is compared to the van Hout et al.’s crosswalk QALE norms on the same lifetable. Variable discount rates were applied at 4%, 3% and 2% as per guidelines.

Based on the newer population norms the QALE of average population has decreased by 4%. The QALE at birth was previous 69.62 years for men and 69.67 years for women which came down to 64.93 years and 68.77 years respectively.

The current study quantifies the diminishing QALEs based on the updated QALE estimates at the same time allowing the estimation of absolute and proportional shortfall.

## Introduction

According to the framework for priority setting established by the third Norwegian Committee on Priority Setting in Health Sector states that prioritization is founded on three statutory criteria; 1) expected health-benefit, 2) resources use, and 3) health-loss [1]. While the first two criteria are operationalized through cost-effectiveness ratios, the third one is constitutes of severity and is quantified by the absolute and proportional quality-adjusted life-year (QALY) shortfall. Shortfall constitutes of the healthy life years lost due to premature death and reduced quality of life during a period of illness, either in absolute or proportion form [2]. However, it is more than just descriptive statistics; they are critical inputs directly influencing the reimbursement decisions.

Historically, Norway lacked a domestic EQ-5D value set and relied on the UK-derived EQ-5D-3L tariff with van Hout crosswalk to 5L data [3,4]. This practice had several well-documented limitations: 1) interim and outdated solution, 2) artificial flooring effect from crosswalk, 3) difference in population and health preferences [5]. To address these limitations, the Norwegian Institute of Public Health and the Norwegian Medical Products Agency commissioned a nationally representative study [3,6]. Using a hybrid composite time trade-off (cTTO) and discrete choice experiment approach, the study produced new tariff ranging from −0.453 to 1, with anxiety/depression emerging as the single most influential dimension [7].

An international comparison study demonstrated the average reduction in health gained to range between 30% to 84% when adapting the 5L tariff from 3L [8]. Hence, transition to the new tariff is not a cosmetic adjustment, and might lead to inflated shortfall estimates and re-rank interventions across set thresholds. In addition to the ageing population of Norway and increased prevalence of multimorbidity, the life expectancy at birth has increased by 2.2 years for men and 1.8 years for women in the past few decades [9,10]. Therefore, up-to-date and accurate quality-adjusted life expectancy (QALE) norms are essential to prevent age-based inequities.

In the current study, QALE norms for ages 0 to 100 by sex are estimated for the Norwegian population using the latest EQ-5D-5L tariff and most recent lifetables (2023-2024). Secondly, these estimates are contrasted against the long-standing 3L tariffs. These hopefully would enable stakeholders to anticipate how the tariff changes will impact with Norway’s severity criteria.

## Materials and methods

### Data and sources

Single age and sex specific life expectancy and death rates were derived from the 2024 national life tables available at Statistics Norway [11].

Garratt et al. [12] collected and reported the nationally representative population norms for the EQ-5D-5L for adults (>=18 years) by sex and education. In addition, utility values from the mapped 5L values by van Hout et al. [4] were also used as comparator tariff.

Discount rates were assigned according to the Direktoratet for Medisnske Produkter (DMP) recommendation which states 4% till the age of 39, 3% for population aged between 40-74, and 2% for the remaining ages [3]. QALEs without discount (0%) are also reported.

The life expectancy estimates were combined with the utility values separately to calculate the QALE norms using the Sullivan method [13]. Finally, the age-, sex- and tariff-specific QALE estimates for the Norwegian adult population were reported.

### Assumptions

We made the following assumption prior to the analysis. Lifetable report the highest age of 106, however we limited the analysis to 100 years starting from 0. Tariff values both old and new were available for age groups, and hence we assumed that it remains the same for each individual age within that group. Since the new tariffs did not include feedback from children or youth (0-18 years) they were assumed to have the same health state as the first group, 18-29 years. The life tables also incorporate half-cycle corrections by assuming that individuals dying at age x, died at the middle of the 6, having lived six months.

### Software

All calculations were done in Excel. However, in addition, to enhance user experience and expand the scope of the project, we developed an R Shiny based interactive application to estimate the absolute and proportion shortfall, switch between tariffs, apply varying discount rates, and modify starting age, as well as proportion of male or female in the population for the five Nordic countries (Denmark, Finland, Norway, Iceland and Sweden). The Nordic Shortfall Calculator is available at https://mohasal.shinyapps.io/nordic-shortfall-calculator/. The R code can be provided upon request.

## Results

The age-specific undiscounted and discounted QALE are reported in Table 1 andTable 2, for females and males respectively. The discount is applied using tiered rates; 4% for ages 0-39, 3% for ages 40-74, and 2% for ages above 75.

**Table 1:**
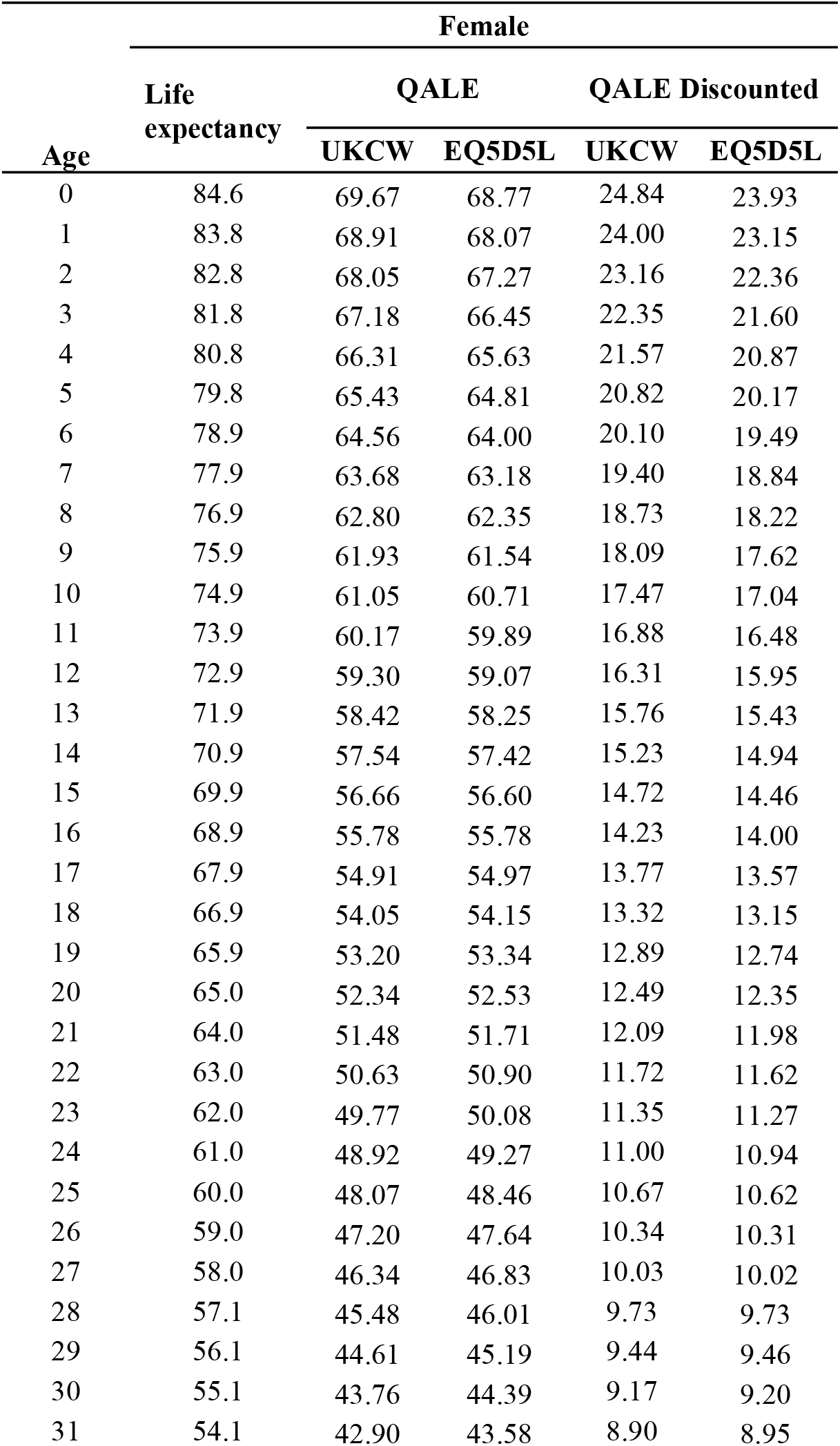
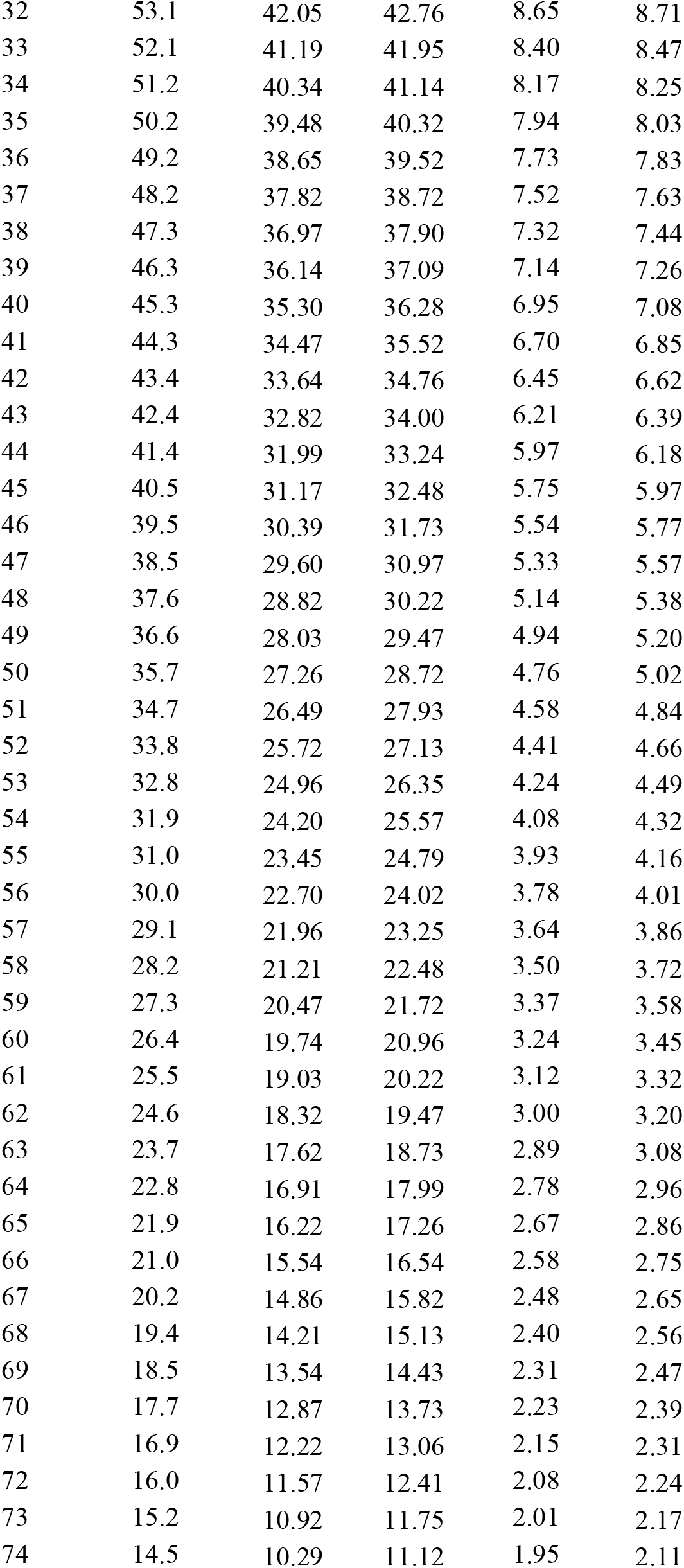
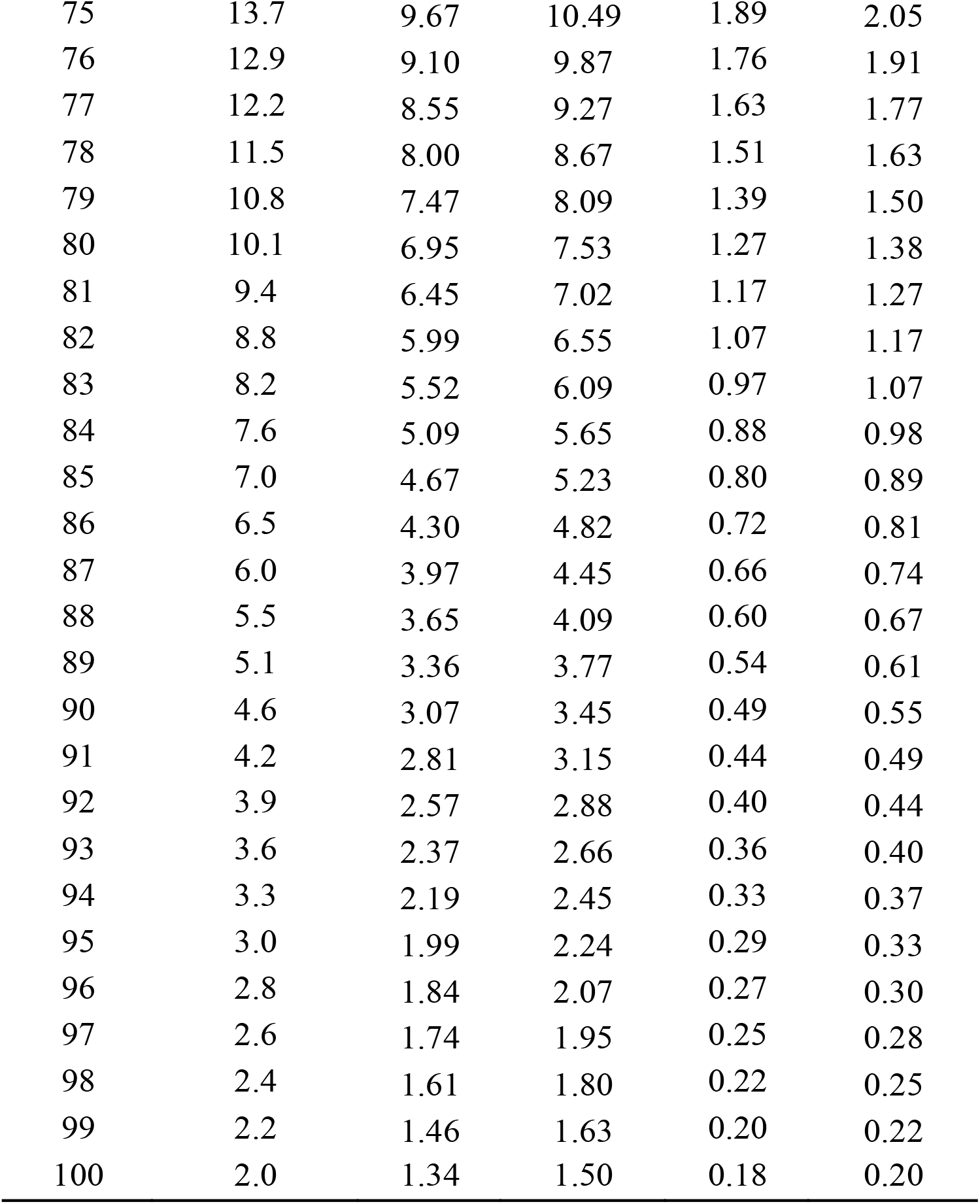
Life expectancy for females according to old and new tariffs without and with discount (4% till age 39, 3% for 40-74, and 2% for 75 and above; according to NOMA)

**Table 2:** Life expectancy for males according to old and new tariffs without and with discount (4% till age 39, 3% for 40-74, and 2% for 75 and above; according to NOMA)

| Age | Male |  |  |  |  |
| --- | --- | --- | --- | --- | --- |
|  | Life expectancy | QALE |  | QALE Discounted |  |
|  |  | UKCW | EQ5D5L | UKCW | EQ5D5L |
| 0 | 81.4 | 69.63 | 64.93 | 22.71 | 20.63 |
| 1 | 80.6 | 68.87 | 64.26 | 21.85 | 19.87 |
| 2 | 79.6 | 67.97 | 63.46 | 20.97 | 19.09 |
| 3 | 78.6 | 67.06 | 62.65 | 20.13 | 18.34 |
| 4 | 77.6 | 66.16 | 61.85 | 19.31 | 17.62 |
| 5 | 76.6 | 65.26 | 61.05 | 18.54 | 16.93 |
| 6 | 75.6 | 64.35 | 60.24 | 17.79 | 16.26 |
| 7 | 74.7 | 63.44 | 59.44 | 17.06 | 15.62 |
| 8 | 73.7 | 62.53 | 58.63 | 16.37 | 15.00 |
| 9 | 72.7 | 61.62 | 57.82 | 15.70 | 14.41 |
| 10 | 71.7 | 60.71 | 57.01 | 15.06 | 13.84 |
| 11 | 70.7 | 59.80 | 56.20 | 14.44 | 13.29 |
| 12 | 69.7 | 58.88 | 55.39 | 13.85 | 12.76 |
| 13 | 68.7 | 57.98 | 54.59 | 13.28 | 12.25 |
| 14 | 67.7 | 57.07 | 53.78 | 12.73 | 11.77 |
| 15 | 66.7 | 56.16 | 52.97 | 12.20 | 11.30 |
| 16 | 65.7 | 55.26 | 52.17 | 11.70 | 10.85 |
| 17 | 64.7 | 54.36 | 51.37 | 11.21 | 10.42 |
| 18 | 63.8 | 53.46 | 50.57 | 10.74 | 10.00 |
| 19 | 62.8 | 52.56 | 49.79 | 10.29 | 9.61 |
| 20 | 61.8 | 51.66 | 49.00 | 9.85 | 9.22 |
| 21 | 60.9 | 50.78 | 48.21 | 9.45 | 8.86 |
| 22 | 59.9 | 49.92 | 47.43 | 9.06 | 8.51 |
| 23 | 58.9 | 49.05 | 46.64 | 8.69 | 8.17 |
| 24 | 58.0 | 48.19 | 45.85 | 8.33 | 7.84 |
| 25 | 57.0 | 47.32 | 45.06 | 7.99 | 7.53 |
| 26 | 56.0 | 46.45 | 44.28 | 7.66 | 7.23 |
| 27 | 55.1 | 45.58 | 43.49 | 7.34 | 6.94 |
| 28 | 54.1 | 44.72 | 42.70 | 7.03 | 6.66 |
| 29 | 53.1 | 43.84 | 41.91 | 6.74 | 6.39 |
| 30 | 52.2 | 42.97 | 41.12 | 6.45 | 6.13 |
| 31 | 51.2 | 42.09 | 40.29 | 6.17 | 5.88 |
| 32 | 50.2 | 41.21 | 39.48 | 5.91 | 5.63 |
| 33 | 49.3 | 40.32 | 38.66 | 5.65 | 5.39 |
| 34 | 48.3 | 39.44 | 37.84 | 5.40 | 5.16 |
| 35 | 47.3 | 38.55 | 37.01 | 5.17 | 4.94 |
| 36 | 46.4 | 37.71 | 36.19 | 4.95 | 4.73 |
| 37 | 45.4 | 36.88 | 35.37 | 4.75 | 4.53 |
| 38 | 44.5 | 36.05 | 34.55 | 4.55 | 4.33 |
| 39 | 43.5 | 35.22 | 33.74 | 4.36 | 4.14 |
| 40 | 42.6 | 34.40 | 32.92 | 4.18 | 3.96 |
| 41 | 41.6 | 33.57 | 32.17 | 4.00 | 3.80 |
| 42 | 40.7 | 32.74 | 31.42 | 3.83 | 3.65 |
| 43 | 39.7 | 31.92 | 30.67 | 3.66 | 3.49 |
| 44 | 38.8 | 31.08 | 29.92 | 3.49 | 3.35 |
| 45 | 37.8 | 30.25 | 29.17 | 3.34 | 3.20 |
| 46 | 36.8 | 29.46 | 28.41 | 3.19 | 3.06 |
| 47 | 35.9 | 28.69 | 27.67 | 3.05 | 2.93 |
| 48 | 35.0 | 27.91 | 26.92 | 2.91 | 2.80 |
| 49 | 34.0 | 27.14 | 26.18 | 2.78 | 2.67 |
| 50 | 33.1 | 26.36 | 25.43 | 2.65 | 2.55 |
| 51 | 32.1 | 25.57 | 24.71 | 2.53 | 2.43 |
| 52 | 31.2 | 24.79 | 24.00 | 2.41 | 2.32 |
| 53 | 30.3 | 24.01 | 23.29 | 2.29 | 2.21 |
| 54 | 29.4 | 23.24 | 22.59 | 2.18 | 2.11 |
| 55 | 28.5 | 22.48 | 21.89 | 2.07 | 2.01 |
| 56 | 27.6 | 21.73 | 21.19 | 1.96 | 1.91 |
| 57 | 26.6 | 20.98 | 20.50 | 1.86 | 1.82 |
| 58 | 25.8 | 20.25 | 19.81 | 1.76 | 1.73 |
| 59 | 24.9 | 19.51 | 19.12 | 1.67 | 1.64 |
| 60 | 24.0 | 18.80 | 18.45 | 1.58 | 1.56 |
| 61 | 23.1 | 18.08 | 17.73 | 1.49 | 1.47 |
| 62 | 22.2 | 17.37 | 17.03 | 1.41 | 1.39 |
| 63 | 21.4 | 16.67 | 16.33 | 1.33 | 1.31 |
| 64 | 20.5 | 15.99 | 15.65 | 1.26 | 1.23 |
| 65 | 19.7 | 15.31 | 14.97 | 1.18 | 1.16 |
| 66 | 18.9 | 14.66 | 14.30 | 1.11 | 1.09 |
| 67 | 18.1 | 14.00 | 13.64 | 1.05 | 1.02 |
| 68 | 17.3 | 13.36 | 12.98 | 0.98 | 0.96 |
| 69 | 16.5 | 12.72 | 12.33 | 0.92 | 0.89 |
| 70 | 15.7 | 12.11 | 11.70 | 0.86 | 0.83 |
| 71 | 14.9 | 11.47 | 11.08 | 0.80 | 0.78 |
| 72 | 14.2 | 10.86 | 10.49 | 0.75 | 0.72 |
| 73 | 13.4 | 10.26 | 9.90 | 0.70 | 0.67 |
| 74 | 12.7 | 9.67 | 9.33 | 0.65 | 0.63 |
| 75 | 12.0 | 9.10 | 8.77 | 0.60 | 0.58 |
| 76 | 11.3 | 8.55 | 8.22 | 0.56 | 0.54 |
| 77 | 10.7 | 8.01 | 7.68 | 0.52 | 0.49 |
| 78 | 10.0 | 7.48 | 7.15 | 0.48 | 0.45 |
| 79 | 9.4 | 6.96 | 6.63 | 0.44 | 0.42 |
| 80 | 8.8 | 6.46 | 6.12 | 0.40 | 0.38 |
| 81 | 8.2 | 5.97 | 5.69 | 0.36 | 0.35 |
| 82 | 7.6 | 5.52 | 5.31 | 0.33 | 0.32 |
| 83 | 7.1 | 5.07 | 4.92 | 0.30 | 0.29 |
| 84 | 6.5 | 4.61 | 4.53 | 0.27 | 0.26 |
| 85 | 6.0 | 4.20 | 4.19 | 0.24 | 0.24 |
| 86 | 5.5 | 3.85 | 3.86 | 0.22 | 0.22 |
| 87 | 5.1 | 3.52 | 3.55 | 0.20 | 0.20 |
| 88 | 4.7 | 3.19 | 3.26 | 0.18 | 0.18 |
| 89 | 4.2 | 2.86 | 2.95 | 0.16 | 0.16 |
| 90 | 3.9 | 2.60 | 2.74 | 0.14 | 0.15 |
| 91 | 3.6 | 2.39 | 2.51 | 0.13 | 0.13 |
| 92 | 3.3 | 2.17 | 2.28 | 0.11 | 0.12 |
| 93 | 3.0 | 1.99 | 2.09 | 0.10 | 0.11 |
| 94 | 2.8 | 1.85 | 1.94 | 0.09 | 0.10 |
| 95 | 2.6 | 1.72 | 1.81 | 0.08 | 0.09 |
| 96 | 2.3 | 1.55 | 1.63 | 0.07 | 0.08 |
| 97 | 2.2 | 1.47 | 1.54 | 0.07 | 0.07 |
| 98 | 2.1 | 1.39 | 1.46 | 0.06 | 0.07 |
| 99 | 1.9 | 1.25 | 1.32 | 0.06 | 0.06 |
| 100 | 1.6 | 1.06 | 1.11 | 0.05 | 0.05 |

At birth, females are expected to live longer than males (84.6 vs 81.4 years), a difference of 3.2 years. However, the gap in QALE varies depending on the valuation method used. Using the UK crosswalk tariff, QALE at birth is near identical for both females and males (69.67 vs. 69.63 QALYs), whereas the newer EQ-5D-5L tariff shows a more pronounced difference (68.77 vs 64.93 QALYs), favoring females by 3.84 QALYs.

Discounting substantially reduced the QALE estimates. At birth, discounted QALE under the UK crosswalk is 24.84 QALYs for females and 22.71 QALYs for males while the EQ-5D-5L yields 23.93 and 20.63 QALYs, respectively. The gender gap in discounted QALE is thus more prominent under the EQ-5D-5L (3.3 QALYs) than the UK crosswalk (2.1 QALYs).

As age increases, both life expectancy and QALE estimates decline steadily for both sexes. Discrepancy in QALE between females and males are more evident under the EQ-5D-5L tariff. At age 60, a difference of 2.51 QALYs is observed under the EQ-5D-5L tariff in contrast to 0.94 QALYs under the UK crosswalk.

## Discussion

Norway’s recent adoption of the EQ-5D-5L value set marks a significant shift in how health-realted quality of life is quantified for health technological assessments. This study provides updated QALE population norms for Norway, stratified by age and sex, using both the newly promoted EQ-5D-5L tariff and the previously used UK crosswalk method. These norms are essential for estimating absolute and proportional shortfall, which are pivotal to Norway’s prioritization framework, also endorsed by DMP [3].

The results demonstrate that while the life expectancy remains higher for females than males, the change in choice of tariff would substantially influence the QALE estimates, and also the magnitude of sex-based difference, thereby subsequently also affecting the absolute and proportional shortfall. While the discrepancy in between the sexes widens under both the tariffs as the age progresses, however it is more prominent under the newer EQ-5D-5L tariff, favoring females over males. The EQ-5D-5L tariff might be better at capturing sex-specific gains in health-related quality of life. The reduction in QALE norms upon adapting the newer tariffs has critical implications on severity assessment, as it directly modifies the absolute and proportional shortfall estimates. This might in turn re-rank intervention and potentially alter reimbursement decisions.

Current findings of reduction in health gains when transitioning from 3L-based tariff to 5L-based aligns well with international findings as well [5,14,15]. The average health-utility state as reported by Norwegians are amongst the lowest globally, indicating towards a more conservative approach while perceiving one’s health states [5]. The newer tariff emphasizes the mental health dimension, which also aligns with the broader trends observed in neighboring countries [7,16– 18]. The cross-country differences mentioned by Wang et. al. [5], emphasizes the importance of using locally derived, culturally sensitive value sets especially when Norway exhibits relatively lower ceiling effects, indicating that fewer individuals report perfect health.

There are several limitations in the current study; firstly, information on health-related quality of life was not collected for population below 18 years of age, secondly, the utility values were applied uniformly between the age groups, which may skew variation, finally, while the Sullivan method provides a robust framework on combining the mortality and morbidity data, it is limited to current population health and does not account for improvement in health of the society in the future.

Despite these limitations, the updated QALE norms presented here offer a more accurate and nationally relevant benchmark for assessing the severity in Norway. Thus it promotes the broader application of EQ-5D-5L scale in clinical effect evaluation, economic evaluation, as well as policy making [19,20]. They also provide a foundation for consistent and transparent application of shortfall metrics in health technology assessments and support the broader goal of equitable prioritization in healthcare.

## Data Availability

All data are available on publicly accessible respective governmental portals.

https://www.ssb.no/

## Acknowledgements

The Nordic Shortfall Calculator has been developed taking inspiration from UK QALY Shortfall (https://shiny.york.ac.uk/shortfall) developed by McNamara et al.

